# Oral-gut microbiome profiles in environmentally matched dizygotic triplets discordant for autism spectrum disorder: an exploratory study

**DOI:** 10.64898/2026.08.14.26360454

**Authors:** Nathália Tuany Duarte, Caroline Augusta Belo Faria, Joyce Vanessa da Silva Fonseca, Franciane Mendes de Oliveira, Ester Cerdeira Sabino, Paulo Henrique Braz da Silva, Fabiana Martins, Marina Gallottini

## Abstract

**Background/Objectives:** Autism spectrum disorder (ASD) has been associated with microbiome alterations, but the relative contribution of environmental and individual factors remains unclear. This study explored oral and gut microbiome profiles in environmentally matched dizygotic triplets discordant for ASD.

**Materials and Methods:** Triplets in the 5–9-year age range, including one child with ASD and two neurotypical siblings, underwent standardized oral examination. Oral tongue-dorsum and rectal swab samples were analyzed by 16S rRNA sequencing. Taxonomic composition and beta diversity were evaluated descriptively.

**Results:** Dominant bacterial phyla were broadly similar across siblings, but oral microbial profiles showed greater interindividual variation. The participant with ASD had the highest dental biofilm accumulation, predominance of *Streptococcus*, and reduced representation of several secondary genera. One neurotypical sibling with mild gingival inflammation showed greater representation of *Fusobacterium*, *Prevotella*, and *Leptotrichia*. Beta diversity demonstrated clearer interindividual separation among oral than gut samples.

**Conclusions:** Individual-specific factors may influence microbiome patterns even under highly similar environmental and dietary conditions. These findings support further investigation of the oral microbiome as a complementary component of ASD microbiome research.

## Introduction

Autism spectrum disorder (ASD) is a neurodevelopmental condition with a complex and multifactorial etiology involving genetic, epigenetic, and environmental factors [1]. Increasing evidence supports a role for the human microbiome in modulating neurobiological processes underlying development and behavior [2,3].

Over the past decades, the gut microbiome has been extensively investigated in ASD. Gastrointestinal symptoms are frequently reported in individuals with ASD, and gut microbial alterations have been reported compared with typically developing individuals [4–7]. Differences in microbial composition and metabolites, including short-chain fatty acids, may influence immunological, metabolic, and neurobiological processes [8]. These interactions form the gut– brain axis, a bidirectional communication system involving neural, immune, and endocrine pathways [2,7].

Although most research has focused on the gut microbiome, the oral microbiome represents a complex ecosystem composed of hundreds of bacterial species that play important roles in oral and systemic health [9,10]. The oral cavity is one of the main entry points to the gastrointestinal tract and may act as a reservoir of microorganisms capable of influencing the gut microbiome [11]. Despite this relevance, the oral microbiome remains underexplored in ASD, particularly in studies that simultaneously consider clinical oral health conditions, such as dental caries, periodontal disease, and oral hygiene, which are known to influence oral microbial composition [12–14].

Studies integrating both oral and gut microbiomes in individuals with ASD may contribute to a more comprehensive understanding of microbial patterns associated with neurodevelopment [15,16]. In this context, highly comparable family groups, such as siblings, twins, or triplets, may help reduce genetic and environmental variability [17–19].

Therefore, the present study aims to describe the composition of the oral and gut microbiomes of dizygotic triplets, one with ASD and two with typical neurodevelopment, to explore potential microbial differences under highly shared environmental conditions.

## Materials and Methods

This exploratory study included three non-monozygotic triplet siblings, in the 5–9-year age range (#1, #2, and #3), living in the same household and exposed to similar environmental conditions. The study was approved by the institutional Research Ethics Committee (protocol number 7.264.721), and written informed consent was obtained from the participants’ legal guardians. One participant (#1) had a diagnosis of autism spectrum disorder (ASD), while the other two (#2 and #3) were typically developing.

The triplets had similar dietary habits, as documented by a three-day food diary, and similar oral hygiene practices. None of the participants had received antibiotic or probiotic therapy in the three months prior to sample collection. All three siblings were clinically evaluated by the same dentists, and intraoral examination was performed under standardized conditions. Oral health parameters included plaque index, gingival index, and caries indices, including the International Caries Detection and Assessment System (ICDAS) and decayed, missing, and filled teeth index (dmft), with all evaluations conducted in a standardized manner to allow comparison among participants. Prior to data collection, examiner calibration for ICDAS assessment was conducted using a reference examiner, demonstrating excellent agreement between the calibrated examiner and the reference standard (κ = 0.923), as well as intraexaminer agreement (κ = 0.921).

Oral samples were collected from the tongue dorsum using sterile swabs by the dentists during clinical evaluation, while rectal swab samples were collected on the same day by a trained nurse. All samples were obtained using sterile swabs and stored at -80 °C until processing. Bacterial genomic DNA was extracted using the DNeasy PowerSoil Pro Kit (QIAGEN, Hilden, Germany), and DNA was quantified by Qubit® 3.0 fluorometer using dsDNA High Sensitivity Assay Kit (Thermo Fisher Scientific, Massachusetts, USA). The hypervariable V4 region of the 16S rRNA gene was amplified using the 515F and 806R primers [20], and sequenced using the Ion GeneStudio S5 system (Thermo Fisher Scientific, Massachusetts, USA).

Sequencing data were processed using QIIME 2 (version 2024.10.1). Quality filtering and amplicon sequence variant (ASV) inference were performed using DADA2. Taxonomic classification was performed using the Greengenes database. Microbial composition was assessed based on relative abundance at phylum and genus levels. Results were visualized using bar plots to allow comparison between individuals and sampling sites (oral and gut). Low-abundance taxa were further explored using bubble plots. Beta diversity was assessed using Bray-Curtis distance metrics. Formal alpha diversity comparisons were not performed due to the small sample size.

This analysis represents a subset of participants from a broader ongoing study investigating the oral and gut microbiome in individuals with ASD and their siblings. Considering the small sample size (n = 3), analyses were conducted in a descriptive and exploratory manner, without inferential statistical testing.

## Results

### Clinical characteristics of participants

Participant #1 (ASD) exhibited the highest dental biofilm accumulation and multiple early-stage caries lesions, without gingival inflammation. Participant #2 showed mild gingival inflammation and a single early-stage caries lesion. Participant #3 presented the lowest biofilm accumulation, no gingival inflammation, and one early-stage lesion. (Table 1)

**Table 1.** Baseline clinical characteristics.

| **Participant** | **Plaque Index** | **Gingivitis** | **Initial Lesions (ICDAS)** | **dmft** |
| --- | --- | --- | --- | --- |
| #1 (ASD) | 0.833 | Absent | A / 3 lesions | 0 |
| #2 | 0.667 | Mild | A / 1 lesion | 0 |
| #3 | 0.333 | Absent | A / 1 lesion | 0 |

dmft: Decayed, Missing, and Filled Teeth; ICDS: International Caries Detection and Assessment System; ASD: Autism spectrum disorder

### Oral microbiome composition

At the phylum level, the oral microbiome showed predominance of Bacillota in all three siblings, with higher relative abundance in participants #1 and #3. The phylum Pseudomonadota was also identified in all individuals, with greater representation in participants #2 and #3 compared to the individual with ASD (#1). Additionally, Bacteroidota showed higher relative abundance in participant #2, while being less represented in participants #1 and #3. Other phyla, such as Fusobacteriota and Actinomycetota, were detected at lower proportions, with interindividual variation. (Figure 1)

**Figure 1.**
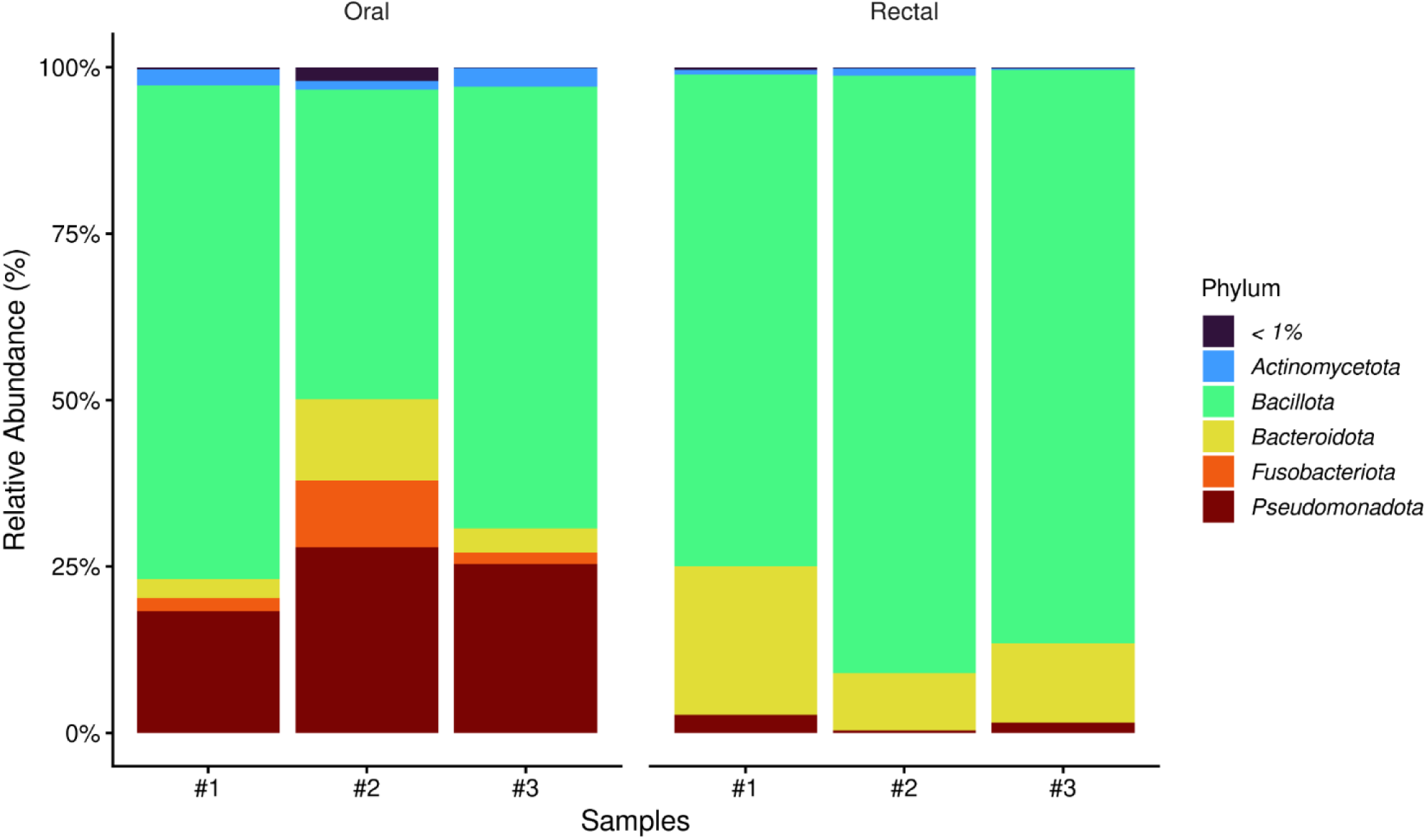
Bar plot showing the relative abundance of oral and gut microbiome at the phylum level across participants.

At the genus level, *Streptococcus* was predominant across all oral samples, with relative abundances of 68.7%, 27.8%, and 53.7% in participants #1, #2, and #3, respectively. In contrast, participant #2 exhibited a more heterogeneous microbial profile, with greater representation of secondary genera, including *Neisseria* (5.2%) and *Granulicatella* (5.4%). Both *Neisseria* and *Granulicatella* were more abundant in the neurotypical siblings, particularly participant #3 (9.5% and 6.8%, respectively), while showing minimal representation in the individual with ASD (#1). (Figure 2)

**Figure 2.**
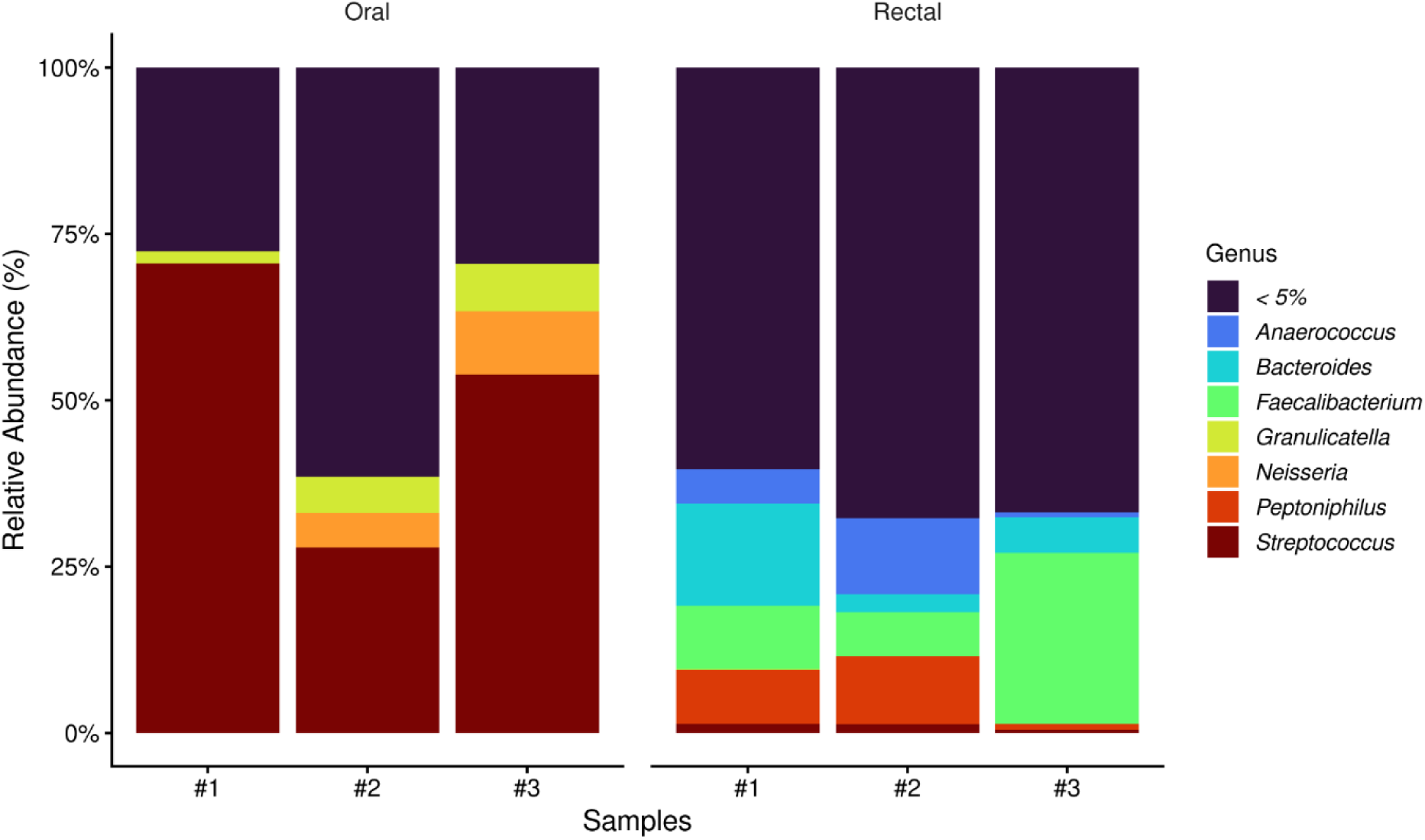
Bar plot showing the relative abundance of oral and gut microbiome at the genus level across participants.

Evaluation of lower-abundance taxa further highlighted interindividual variation in microbial organization. The participant with ASD (#1) exhibited reduced representation of several secondary genera, including *Veillonella*, which showed relative abundances of 3.4% and 0.5% in neurotypical participants #2 and #3, respectively, while being minimally represented in participant #1. In contrast, the neurotypical siblings exhibited broader representation of secondary genera within the oral microbiome. (Figure 3)

**Figure 3.**
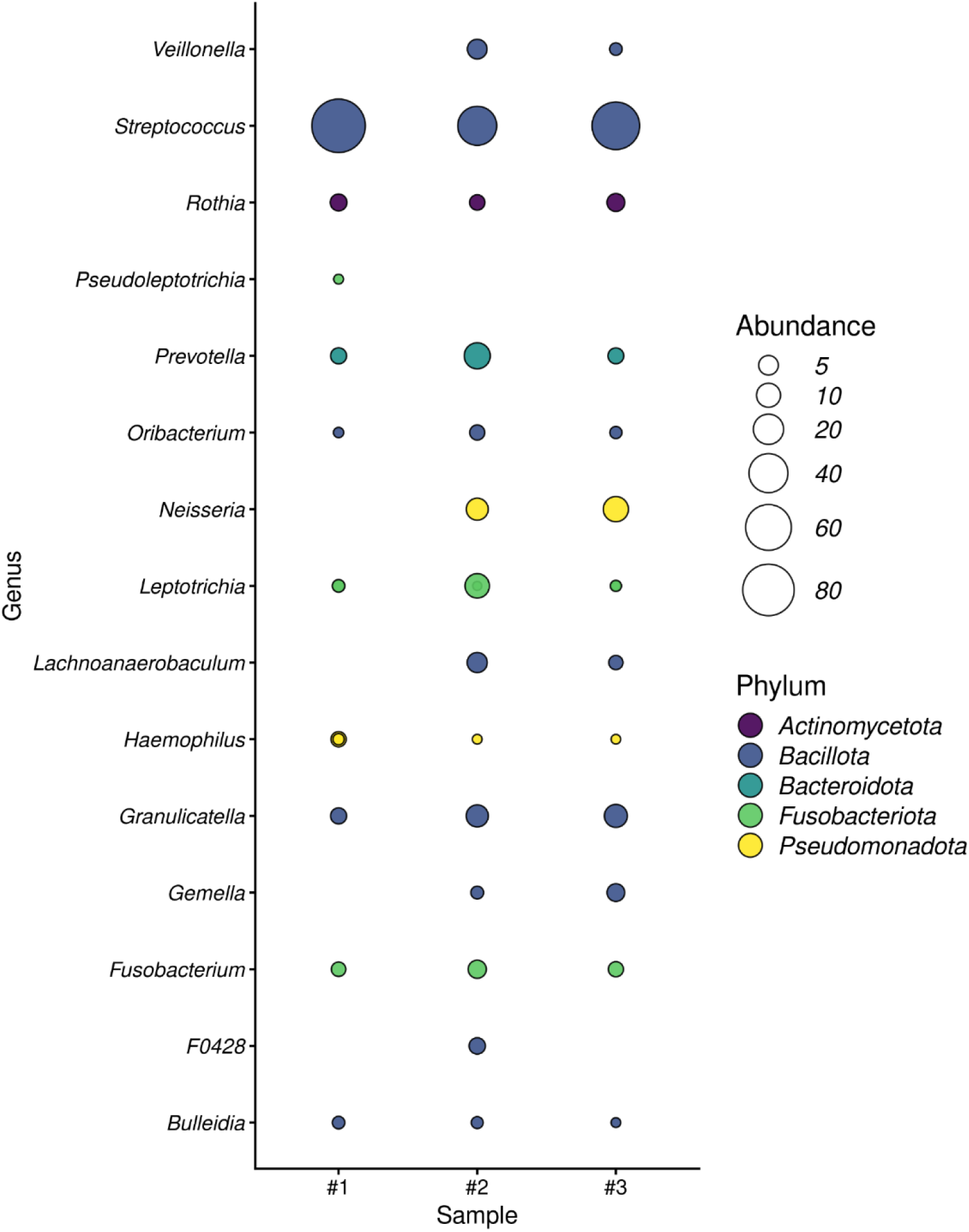
Distribution of low-abundance oral bacterial genera across triplet siblings. Note: Bubble size represents the relative abundance of each bacterial genus, while colors indicate taxonomic classification at the phylum level. This visualization highlights the distribution of low-abundance taxa and interindividual variation in oral microbiome composition.

Participant #2 exhibited greater representation of genera such as *Fusobacterium* (2.6%), *Prevotella* (2.8%), and *Leptotrichia* (5.6%). In participants #1 and #3, *Fusobacterium* showed relative abundances of 1.0% and 1.4%, *Prevotella* of 0.4% and 0.1%, and *Leptotrichia* of 0.2% and 0.1%, respectively. Participant #2 was also the only individual presenting gingival inflammation. In contrast, participant #3 exhibited lower representation of these genera.

### Gut microbiome composition

Analysis of the gut microbiome revealed predominance of the phyla Bacillota and Bacteroidota across all samples, with variations in their relative proportions among individuals. (Figure 1)

At the genus level, bacteria associated with short-chain fatty acid production, such as *Faecalibacterium* (9.5% and 25.7%) and *Bacteroides* (6.3% and 3.4%), were observed with higher abundance in participants #1 and #3, respectively. (Figure 2)

Analysis of low-abundance taxa revealed interindividual differences in gut microbiome composition. Participant #2 exhibited greater representation of genera such as *Staphylococcus*, *Finegoldia*, and *Anaerococcus* compared to the other siblings, whereas participant #3 showed lower representation of these microorganisms. (Figure 4)

**Figure 4.**
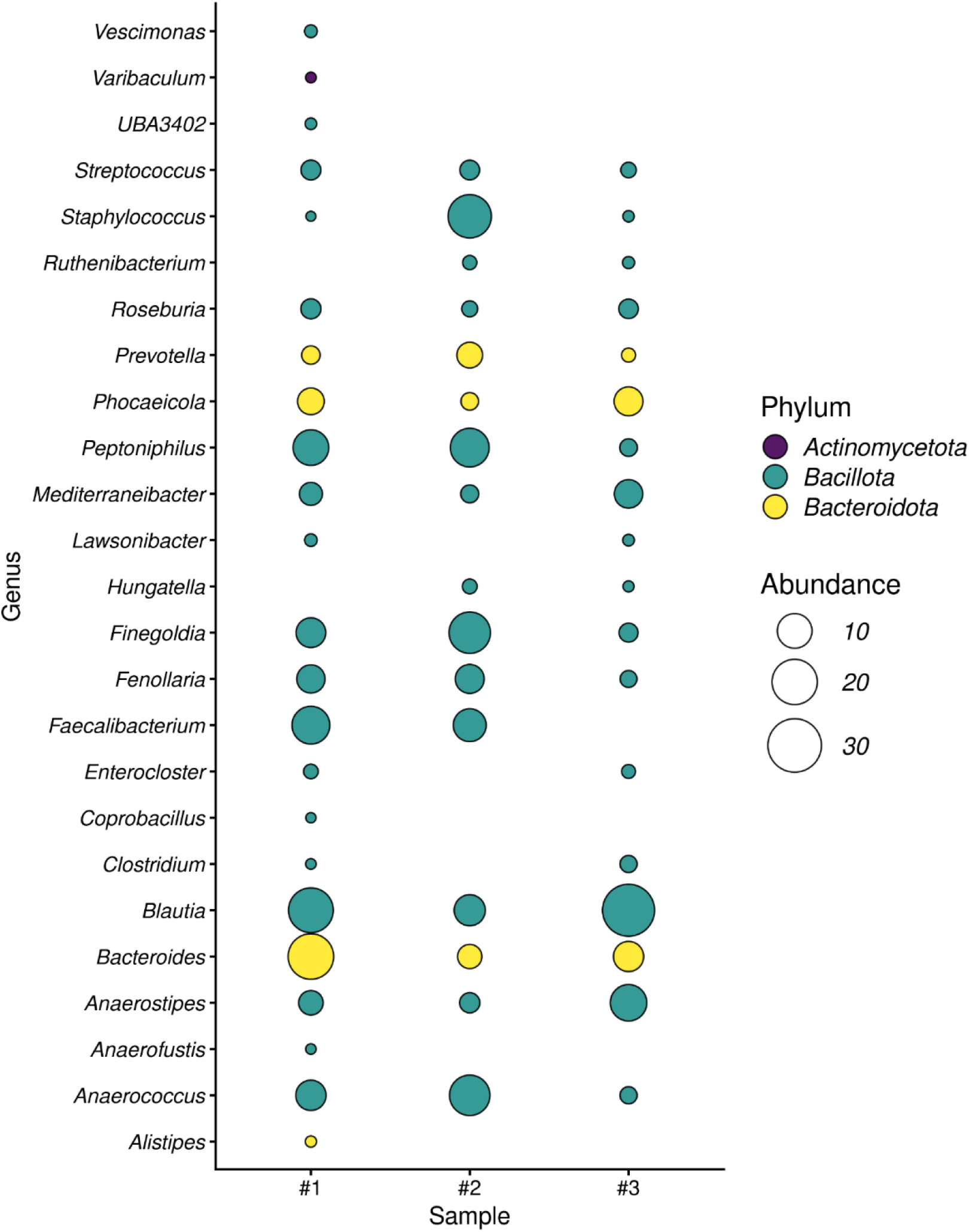
Distribution of low-abundance gut bacterial genera across triplet siblings. Note: Bubble size represents the relative abundance of each bacterial genus, and colors correspond to bacterial phyla. The plot emphasizes differences in low-abundance taxa among individuals, revealing interindividual variability in gut microbiome composition.

### Beta diversity analysis

Beta diversity was assessed using Principal Coordinates Analysis (PCoA) based on the Bray-Curtis distance matrix. (Figure 5)

**Figure 5.**
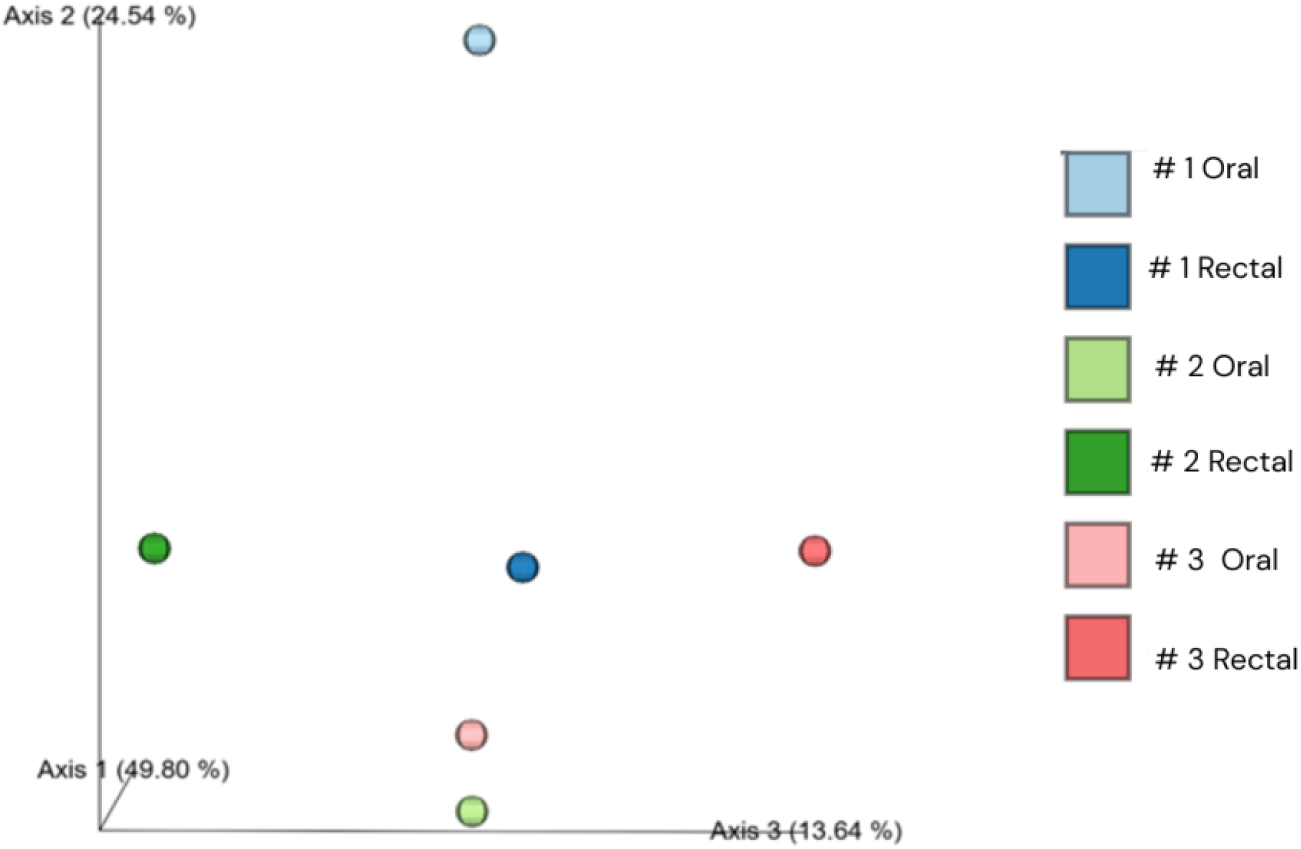
Principal Coordinates Analysis (PCoA) based on Bray–Curtis distances showing the distribution of oral and rectal microbiome samples across triplet siblings. Note: Each point represents an individual sample. Colors indicate participant identity (#1, #2, and #3), and shading distinguishes sample origin (oral vs. rectal). The first two axes explain 49.80% and 24.54% of the total variance, respectively. Separation between oral and rectal samples indicates strong site-specific differences in microbial community structure.

The first two axes explained a substantial proportion of data variability (Axis 1: 49.80%; Axis 2: 24.54%), accounting for approximately 74% of the observed variation. A clear overall separation between oral and gut samples was observed, indicating that anatomical site strongly influences microbial community structure. Oral and rectal samples occupied distinct regions in multivariate space, although some samples showed partial proximity in the ordination plot.

When analyzed by individual, oral microbiome samples exhibited greater interindividual variation than gut samples, despite shared environmental and dietary conditions. Participant #1, diagnosed with ASD, showed a more distinct oral microbiome profile compared to the neurotypical siblings, as evidenced by the separation of the oral sample in multivariate space. In contrast, gut microbiome samples appeared more closely distributed among the three participants.

## Discussion

The findings of this study showed overall separation between oral and gut samples in the beta diversity analysis, alongside interindividual variation in microbial profiles. In triplets with shared environmental and dietary conditions, these differences suggest a contribution of individual-specific factors to microbiome organization, as previously described [18].

Participant #3 exhibited lower biofilm accumulation, absence of gingival inflammation, and lower representation of genera associated with oral biofilm maturation, particularly *Fusobacterium*, *Prevotella*, and *Leptotrichia*, compared with participant #2. In gut samples, genera such as *Faecalibacterium* and *Bacteroides*, associated with short-chain fatty acid (SCFA) production and intestinal homeostasis, were observed in participants #1 and #3 [21,22].

A different pattern was observed in participant #2, who presented mild gingival inflammation accompanied by greater representation of genera associated with oral biofilm maturation, particularly *Fusobacterium*, *Prevotella*, and *Leptotrichia*. *Fusobacterium* is an important bridge organism in oral biofilm development, facilitating coaggregation between early and late colonizers [23]. *Prevotella* and *Leptotrichia* have also been associated with anaerobic oral communities and gingival inflammation and biofilm maturation [10].

The participant with ASD (#1), in turn, showed greater dental biofilm accumulation and more early caries lesions, suggesting an oral environment favoring acidogenic microorganisms, particularly *Streptococcus* [24]. The reduced representation of secondary genera such as *Neisseria*, *Granulicatella*, and *Veillonella* suggests a less heterogeneous oral microbial profile compared with the neurotypical siblings. Beta diversity analysis demonstrated a more distinct oral microbiome profile in this participant, consistent with the greater interindividual variation among oral samples. Considering the shared environmental and dietary conditions, these findings suggest that host-related and behavioral factors may contribute to oral microbiome organization in ASD.

In individuals with ASD, several studies have reported alterations in gut microbiome composition, including differences in the abundance of phyla such as *Bacteroidota*, *Bacillota*, and *Actinobacteriota*, as well as genera such as *Bacteroides*, *Clostridium*, and *Prevotella* [25,26]. Changes in short-chain fatty acid–producing bacteria, including *Faecalibacterium*, have also been associated with immunological and neurobiological modulation [2,16]. In the present study, however, interindividual differences were more evident in the oral than gut microbiome, particularly in the participant with ASD. These findings suggest that microbiome-related alterations in ASD may involve distinct patterns across niches, with the oral microbiome representing an important and still underexplored component.

Taken together, these findings suggest that individual-specific factors may influence microbial composition despite highly shared environmental conditions. Moreover, oral microorganisms may interact with gastrointestinal microbial ecosystems through continuous swallowing and cross-niche exposure [11].

Although oral hygiene was performed by the caregiver, the participant with ASD showed lower cooperation during toothbrushing, which may have contributed to increased biofilm accumulation and influenced the oral microbiome. This behavioral difference may act as an indirect modulator of microbial composition by influencing local ecological conditions, highlighting the importance of behavioral context when interpreting microbiome patterns in neurodevelopmental conditions [14].

The main limitations are the small sample size, which restricted analyses to a descriptive and exploratory approach without inferential statistical testing. Although the shared household, similar dietary habits, and similar environmental exposures minimized confounding factors, genetic and behavioral differences may have contributed to the observed microbial patterns. 16S rRNA gene sequencing did not enable functional metagenomic or metabolomic profiling, while the cross-sectional design precludes assessment of temporal stability or causality. Thus, these findings are hypothesis-generating rather than definitive evidence of ASD-specific microbiome alterations.

Overall, these findings support the microbiome as a dynamic and interconnected system shaped by clinical, behavioral, and host factors. By integrating oral and gut microbiome analyses within a shared familial context, this study extends perspectives beyond gut-centered models, highlighting the importance of considering distinct microbial niches in ASD.

## Conclusion

In conclusion, this exploratory study demonstrated distinct oral and gut microbiome patterns among environmentally matched triplet siblings, including increased dental biofilm accumulation and a more distinct oral microbiome profile in the participant with ASD. These findings highlight the influence of individual-specific factors, including behavioral and intrinsic host characteristics, on microbial variability despite highly similar environmental and dietary conditions.

Although limited by the small sample size, this study provides hypothesis-generating evidence supporting the relevance of integrated oral and gut microbiome analyses in ASD research and reinforces the value of combined clinical and microbiological approaches for personalized prevention and oral healthcare strategies.

## Ethical Approval

This study was approved by the local Research Ethics Committee. Written informed consent was obtained from the participants’ legal guardians.

## Acknowledgements

This study was supported by the Next-Generation Sequencing Facility of the University of São Paulo School of Medicine (GEF02 – Rede Premium).

## Author Contributions

Nathália Tuany Duarte: Conceptualization, Methodology, Data curation, Formal analysis, Writing – original draft, Writing – review & editing; Caroline Augusta Belo Faria: Methodology, Data curation, Formal analysis, Writing – review & editing; Joyce Vanessa da Silva Fonseca: Visualization, Formal analysis, Writing – review & editing; Franciane Mendes de Oliveira: Investigation, Formal analysis, Writing – review & editing; Ester Cerdeira Sabino: Supervision, Writing – review & editing; Fabiana Martins: Validation, Writing – review & editing; Paulo Henrique Braz da Silva: Validation, Writing – review & editing; Marina Gallottini: Supervision, Methodology, Project administration, Validation, Writing – review & editing.

## Data availability statement

The data that support the findings of this study are available from the corresponding author upon reasonable request.

## Declaration on generative AI and AI-assisted technologies in the writing process

During the preparation of this work, the authors used ChatGPT (OpenAI) to assist with language refinement and editing of the manuscript. The authors reviewed and approved the final version of the manuscript and take full responsibility for its content.

